# Meal Protein Quality Score: A novel tool to evaluate protein quantity and quality of meals

**DOI:** 10.1101/2024.06.04.24308419

**Authors:** Pol Grootswagers, Sine Højlund Christensen, Marielle Timmer, William Riley, Lisette de Groot, Inge Tetens

## Abstract

**Background:** The recent shift towards increased plant-based protein consumption has necessitated the development of new tools to evaluate the quality and quantity of protein in meals, especially given the changing dietary guidelines and the adoption of plant-centric menus in healthcare and other settings.

**Objective:** To develop and test the feasibility of the Meal Protein Quality Score (MPQS), a novel metric that assesses the protein quality and quantity in meals based on essential amino acid (EAA) content, digestibility and requirements, with a focus on optimizing protein intake for vulnerable populations, particularly older adults.

**Methods:** The MPQS integrates digestibility-adjusted EAA intake with total protein consumed in a meal, that together with the EAA requirements provide a score from 0 to 100 to reflect EAA coverage adequacy. The score was tested for feasibility by applying it to recipe data from real life hospital meals, and to dietary data from the NU-AGE trial, involving detailed seven-day food records from 252 non-vegan participants analyzed over multiple meal moments.

**Results:** The analyses revealed that the higher the content of plant-protein in a meal, the lower the meal protein quality. Also, breakfast meals scored lowest on protein quality, mainly due to low contents of protein overall, and of lysine and methionine. The MPQS effectively highlighted the difference in protein quality between plant-based and animal-based meals, and across different meal types.

**Conclusion:** The MPQS appears to be a practical tool that facilitates the assessment of meal-based protein quality. The MPQS can be used to guide dietary transitions towards plant-rich diets, ensuring that such shifts do not compromise protein adequacy for at-risk populations. The score allows for guidance in meal-planning, leading to improvements in plant-rich meal formulation to meet both individual and public health nutritional needs.

## Introduction

The shift in dietary protein intake towards more plant-based proteins instead of animal-based proteins is gaining traction in consumers, employees, hospital patients and dietary guidelines ^9^. While the beneficial effects of this transition on cardiometabolic outcomes and on environmental sustainability are much welcome, it does pose a health risk on some groups of consumers ^23^. These are in general consumers with increased protein requirements, lower food intakes, at risk of malnutrition or at risk of sarcopenia, such as older adults and patients ^6^. That risk stems from the lower anabolic properties of plant-based proteins, which are a result of the lower protein concentrations and quality.

Protein quality is a product of the digestibility, the essential amino acid contents of a protein source, and the amino acid requirement of the individual. In a diet high in animal protein, protein quality is almost never an issue ^14^. Animal based protein sources typically contain all essential amino acids in proportions similar to our bodily proteins and are generally more than 95% digestible. Hence, in typical Western dietary patterns where 60% of all proteins are consumed through animal sources ^30^, information on digestibility and amino acid contents was not considered crucial for human health, which explains the paucity in these data. However, official dietary guidelines are shifting to more plant-centered diets, and hospitals and meal services are more frequently offering plant-based meals to their consumers, creating a need for data and scoring algorithms to assess the protein quality of meals ^4^.

Calculating protein quality is not only important to plan meals, monitor the current intake, but also to formulate dietary advice to improve protein quality. The current Dutch recommendation for vegans to ensure adequate intake of all essential amino acids is to increase their protein intake with 30% above the recommendation for the general population ^9^. For older adults, such an increased intake is challenging, as they are already encouraged to increase their protein intake and while being frequently faced with losses of appetite ^6^. Also, for environmental reasons that may underlie a shift to more plant-based foods, the advice to eat more is counterintuitive.

A more elegant solution to meet EAA requirements is to combine complementary plant-based protein sources, so that they together deliver all amino acids required by the body. When meals are constructed by taking into account the essential amino acid contents and digestibility of the meal components, a protein quality equal to that of an animal based meal can be achieved without significantly increasing the portion size ^13^. The variation in essential amino acid profiles in plants is large, with some plant sources like pea contain significantly more lysine than the reference pattern and less methionine, while others like rice show the opposite trend ^10^. That variation opens the possibility to mix and match sources that together deliver all essential amino acids that should be present in one meal. With data on amino acid profiles of all protein containing foods and their digestibility, together with data on amino acid requirements, and an algorithm to calculate the protein quality, many combinations of multiple plant sources that result in a high protein quality meal can be identified.

Currently, protein quality is calculated by PDCAAS or DIAAS. These scores take into account digestibility and amino acid patterns, and are useful in determining the quality of single protein sources. However, protein sources are rarely eaten individually, but normally as part of a meal with multiple protein sources ^3^. Moreover, a PDCAAS or DIAAS score does not take into account protein quantity, while having a very low intake of a high-quality source can be physiologically meaningless, as it might still be unable to meet metabolic demands. Therefore, a score is needed that reflects both the quality and the quantity of all proteins consumed together within one meal.

In this paper, we present the development and feasibility testing of the Meal Protein Quality Score (MPQS), which includes protein quantity, targeted essential amino acid requirements, and digestibility-adjusted amino acid intakes in a score.

## Methods

### Database development

For the purpose of facilitating protein quality assessments based on Dutch dietary intake data, the food table NEVO ^19^ was augmented to include amino acid profiles and protein digestibility data for all food items containing more than 1% protein. When available, ileal digestibility data were given precedence over fecal values, and data derived from human studies were prioritized over those obtained from pig and rat studies. Furthermore, digestibility data from in vitro models were not considered. The full procedure of this food table extension has been published before ^11^.

### Personalized EAA requirements

The MPQS takes into account personalized requirements for essential amino acids that a meal should deliver to optimally meet the body’s metabolic demand, which is based on a combination of a protein quantity requirement per meal and the amino acid reference patterns set by WHO ^31^.

For protein quantity, we use a requirement of 0.3 gram per kilogram body weight per meal moment, based on studies that show that this amount is sufficient to stimulate muscle protein synthesis ^32^, and, with three main meals and snacks, will result in a total daily intake around 1.0-1.2 g/kg/d which is in line with official recommendations for older adults ^2^ ^22^.

With this protein quantity, we have a target protein intake in grams, which we multiply by the reference amino acid patterns set by FAO/WHO ^31^ expressed in mg EAA per g protein, resulting in the requirements presented in **Table 1**.

**Table 1.**
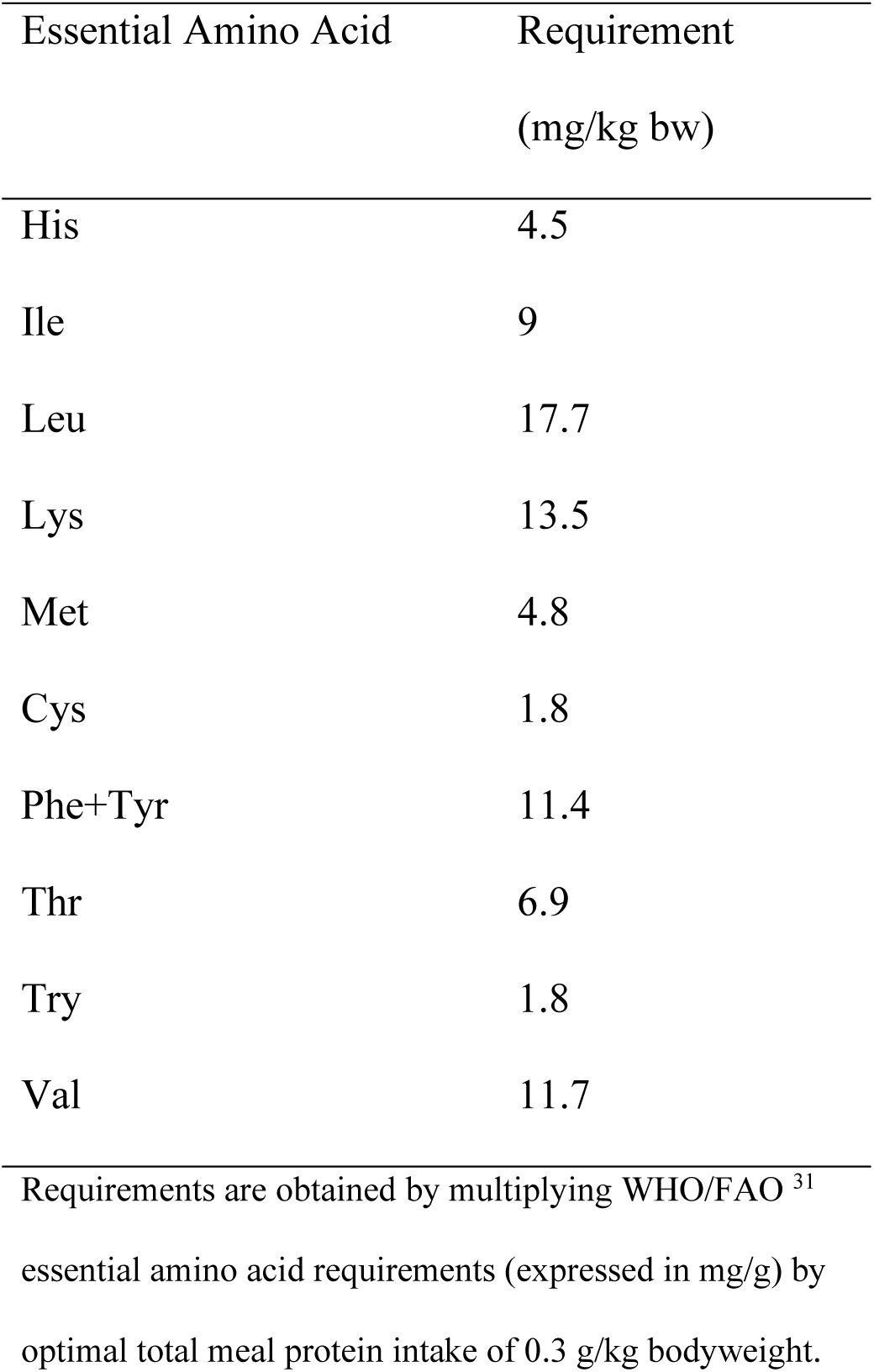
Essential Amino Acid Requirements used to calculate personalized requirements.

| Essential Amino Acid | Requirement<br>(mg/kg bw) |
| --- | --- |
| His | 4.5 |
| Ile | 9 |
| Leu | 17.7 |
| Lys | 13.5 |
| Met | 4.8 |
| Cys | 1.8 |
| Phe+Tyr | 11.4 |
| Thr | 6.9 |
| Try | 1.8 |
| Val | 11.7 |

Requirements are obtained by multiplying WHO/FAO <sup>31</sup> essential amino acid requirements (expressed in mg/g) by optimal total meal protein intake of 0.3 g/kg bodyweight.

### Meal Protein Quality Score

Subsequently, we can score digestibility-adjusted amino acid intakes from a meal. The Meal Protein Quality Score (MPQS) is a composite score that assess protein quality and protein quantity from a meal by taking into account amount of protein, digestibility of protein, and amino acid requirement. MPQS can have values between 0 (where at least one essential amino acid is completely missing in the meal) and 100 (where all essential amino acids reach the requirement). MPQS scores can score above 100 when each amino acid in the meal exceeds the requirement.

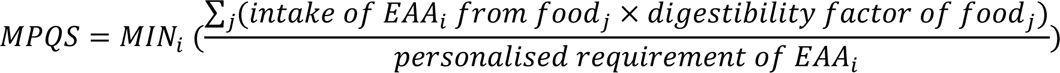

As an example, **Table 2** presents a meal that has an MPQS of 44, meaning that the limiting EAA meets 44% of the requirement. In this case, the digestibility-adjusted intake of methionine is 148 mg, where the meal requirement for this person of 70 kilograms is (70*4.8) 336 mg.

**Table 2.** Example calculation of Meal Protein Quality Score (MPQS). In this meal, the MPQS is 44 and the limiting essential amino acid is methionine.

|  | Digestibility | His | Ile | Leu | Lys | Met | Cys<sup>a</sup> | Phe+<br>Tyr | Thr | Try |
| --- | --- | --- | --- | --- | --- | --- | --- | --- | --- | --- |
| Ingredient A | 94% | 204 | 368 | 628 | 517 | 102 | 107 | 653 | 312 | 103 |
| Ingredient B | 75% | 30 | 62 | 91 | 97 | 23 | 14 | 99 | 56 | 26 |
| Ingredient C | 64% | 29 | 58 | 84 | 92 | 22 | 6 | 73 | 63 | 17 + |
| **Total Digestibility-Adjusted EAA Intake** |  | **263** | **488** | **803** | **706** | **148** | **128** | **825** | **430** | **146** |
| Personalized Meal EAA Requirement<sup>b</sup> |  | 315 | 630 | 1239 | 945 | 336 | 126 | 798 | 483 | 126 ÷ |
| Requirement Met |  | 83% | 77% | 65% | 75% | **44%** | 101% | 103% | 89% | 116% |

<sup>a</sup>Cysteine is topped up with all methionine consumed above the requirement, since methionine can be unidirectionally converted into cysteine. All amino acid intakes are adjusted for the shown digestibility factor before they are summed. <sup>b</sup>In this example, a bodyweight of 70 kilograms is used.

### Application of the score

The practical application of the score was tested for recipes and for large epidemiological datasets. First, recipes of meals that were provided by a hospital in the region of Copenhagen, Denmark, were calculated for protein quality by applying MPQS. The selected meals were developed to contain a higher-than-conventional proportion of plant protein.

Second, the functionality of the MPQS, the score was calculated for all meals consumed at baseline by the Dutch participants of the NU-AGE trial^1^. The participants (n=252) of the NU_AGE trial filled out 7 day food records, resulting in available data on 5121 meals eaten on 1757 days. The NU-AGE trial was a 1-year intervention aimed at improving the diet towards a more Mediterranean diet and had a high-quality dietary assessment^1^. Trained dietitians and nutritionists reviewed food records during home visits, ensuring completeness and accuracy through discussions with participants. Nutrients were calculated by using the NEVO food composition table (NEderlands VOedingsstoffenbestand 2016/5.0 ^19^).

For all meals, data on MPQS, limiting amino acid, (total, animal, and plant) protein intake, and energy intake were calculated.

### Statistical methods

All presented analyses were pre-specified. Descriptive statistics are presented as means ± SD for normally distributed data, and medians with interquartile ranges [IQR; 25th percentile-75th percentile] for non-normally distributed data. The normality of the distribution was visually inspected for all variables, and analyses were conducted using methods appropriate to each distribution type. Differences between groups were evaluated using Analysis of Variance (ANOVA) followed by Tukey’s post hoc test to identify specific differences between meal moments and types. For paired comparisons, such as assessing the impact of adjustments for digestibility, the Wilcoxon signed-rank test was applied. Linear regression analysis was utilized to explore the predictive value of meal characteristics on the MPQS, while Spearman’s rank correlation was employed to develop a correlation matrix. All statistical analyses were conducted using SAS software, version 9.4 (SAS Institute Inc., Cary, NC, USA), and GraphPad Prism, version 9.3.1 (GraphPad Software, San Diego, CA, USA).

## Results

### Applying MPQS to assess protein quality of recipes

A total of 22 real life hospital meals were analysed (**Table 3**). The meals were vegetarian or traditional and had proportions of plant protein varying from 15% to 69%. In meals with plant protein portions below 50%, either leucine or no essential amino acid was limiting. In meals with greater proportions of plant protein, lysine and methionine were limiting.

**Table 3.** Meal Protein Quality Score of 22 real-world hospital meal recipes, sorted by plant portion proportion.

| Recipe name | Portion size (g) | Plant / animal (mg AA /mg AA) | %AA from plant | MPQS | Limiting EAA |
| --- | --- | --- | --- | --- | --- |
| Focaccia with potato, and chanterelle lasagna | 252 | 2908/1325 | 69 | 56 | Methionine |
| Potatoes, and oxheart cabbage | 241 | 1602/925 | 63 | 33 | Methionine |
| Cauliflower soup, rye bread, potato, cream cheese | 221 | 2345/2226 | 51 | 56 | Methionine |
| Mini Danish pastry with cream cheese | 81 | 774/772 | 50 | 18 | Lysine |
| Tartlets, rye bread, eggs | 262 | 2755/2839 | 49 | 75 | Leucine |
| Mushroom gnocchi, cod roe, rye bread, cream cheese | 338 | 3795/4998 | 43 | 116 | - |
| White bread, salad, prawns, walnut- mushroom pâté | 253 | 2267/3719 | 38 | 81 | Leucine |
| Sponge cake | 144 | 1391/2365 | 37 | 51 | Leucine |
| Asparagus soup, prawns, and rye bread with potatoes | 212 | 1750/3710 | 32 | 73 | Leucine |
| Greek yogurt, granola, scrambled egg with tofu, and brioche with rhubarb | 240 | 1926/4559 | 30 | 89 | Leucine |
| Tartlets, smoked chicken, and rye bread with eggs | 325 | 2755/7770 | 26 | 137 | - |
| Focaccia with potato, chanterelle lasagna, salmon | 319 | 2908/8183 | 26 | 149 | - |
| Cauliflower soup, fish terrine, rye bread with potatoes and cream cheese | 267 | 2345/6575 | 26 | 120 | - |
| Pommes Macaire, ham, and with Ingrid peas salad | 287 | 1849/5337 | 26 | 92 | Leucine |
| Rye bread with egg, potatoes, and salmon | 197 | 1834/5299 | 26 | 91 | Leucine |
| Macaroni with cheese, ham, and vegetarian meatballs | 241 | 2002/6114 | 25 | 106 | - |
| Lupin beans with rice, chicken, and vegetarian meatballs in curry | 209 | 2071/6895 | 23 | 116 | - |
| Potatoes, roasted beef, and oxheart cabbage | 281 | 1602/5711 | 22 | 97 | Leucine |
| Cannelloni, vegetarian Bolognese, and Bresaola | 319 | 2240/7789 | 22 | 136 | - |
| Mashed potatoes with broad beans, ham, and stew | 250 | 1679/6170 | 21 | 90 | Cysteine |
| Spinach and parmesan egg, bun with cheese, muesli | 251 | 2090/8826 | 19 | 151 | - |
| Leek pie, salmon, and rye bread with eggs | 269 | 2008/11453 | 15 | 180 | - |

AA, Amino Acid; EAA, Essential Amino Acid; MPQS, Meal Protein Quality Score.

### Applying MPQS to assess protein quality in a large dataset

Data from n=252 participants were used, of which 56% were female, with a mean age of 71 ± 4 years (**Table 4**). The overall protein consumption was 1.0 ± 0.25 g/kg/d, with plant-based protein accounting for 40 ± 8% of the total protein intake. When comparing participants based on their MPQS scores, we identified a subset of 102 individuals who achieved the an MPQS of 100 (indicating meeting all meal EAA requirements) in over half of their meals. This subgroup demonstrated similar demographic and physical activity levels to those who did not meet this criterion. Notable differences were observed in terms of body weight, and dietary intake, with the group scoring lower on the MPQS exhibiting higher body weights and consuming less protein and energy overall.

**Table 4.** Baseline characteristics.

|  | Total sample<br>(n=252) | Median MPQS<br><100 (n=150) | Median MPQS<br>≥100 (n=102) |
| --- | --- | --- | --- |
| Gender, % female | 56 | 56 | 55 |
| Age, y | 71 ± 4 | 71 ± 4 | 71 ± 4 |
| Height, cm | 169 ± 8 | 170 ± 8 | 168 ± 8 |
| Weight, kg | 75 ± 13 | 77 ± 15 | 71 ± 10 |
| BMI, kg/m<sup>2</sup> | 26.0 ± 3.6 | 26.8 ± 4.0 | 25.0 ± 2.7 |
| PASE score | 137 ± 53 | 136 ± 54 | 136 ± 54 |
| Protein intake, g/d | 76 ± 16 | 69 ± 13 | 86 ± 15 |
| Protein intake, g/kg/d | 1.0 ± 0.25 | 0.92 ± 0.19 | 1.23 ± 0.23 |
| Of which plant protein, % | 40 ± 8 | 41 ± 9 | 39 ± 8 |
| Energy intake, kcal/d | 1908 ± 411 | 1800 ± 366 | 2070 ± 422 |
| Percentage of meals > 100 MPQS | 44 ± 18 | 33 ± 11 | 61 ± 12 |
| Median MPQS score | 91 [71-112] | 74 [61-87] | 119 [107-134] |

MPQS, Meal Protein Quality Score; PASE, Physical Activity Scale for the Elderly

### Meal Protein Quality Score per main meal moment

**Figure 1** shows the mean MPQS scores per main meal moment, before and after adjustment for digestibility. Clearly, MPQS increases over the meal moments, with the lowest score (at breakfast (57 ± 1) and the highest at dinner (149 ± 2). Digestibility adjustment significantly lowered the MPQS in all meal moments (median decrease in MPQS of 4 (6.5%), 8 (6.3%) and 17 (10.1%) for breakfast, lunch and dinner, respectively). Most frequent limiting EAAs at breakfast were lysine (73%) and methionine (18%, Table 5).

**Figure 1.**
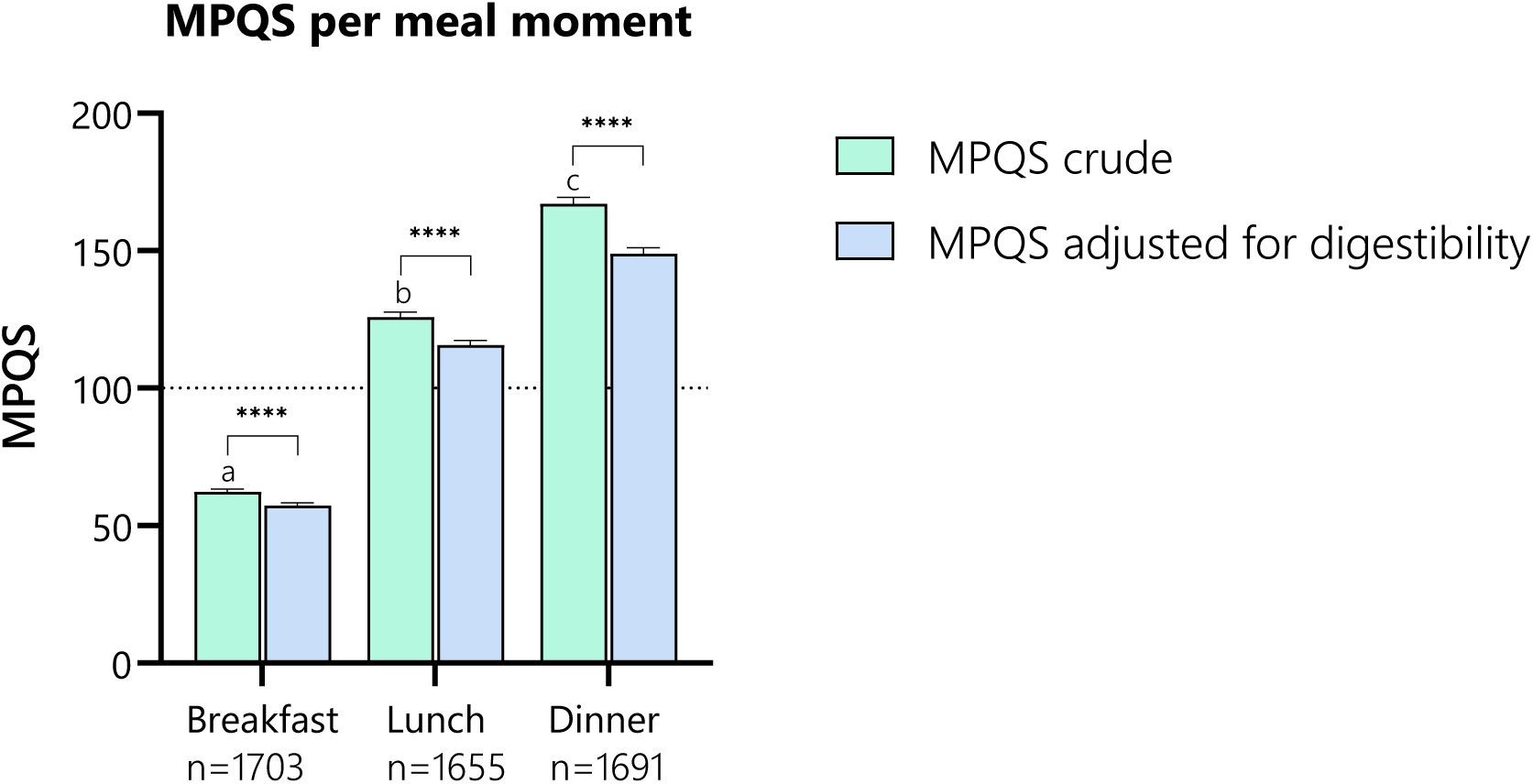
Meal Protein Quality Score by meal moment

**Table 5.** Frequency (%) of limiting essential amino acid per meal moment and type.

|  | Breakfast | Lunch | Dinner | Plant protein<br>0-27% | Plant protein<br>27-65% | Plant protein<br>65-99% | Plant protein<br>100% |
| --- | --- | --- | --- | --- | --- | --- | --- |
| Cysteine | 8 | 11 | 18 | 42 | 8 | 1 | 9 |
| Leucine | 2 | 10 | 27 | 22 | 10 | 2 | 1 |
| Lysine | 73 | 61 | 36 | 4 | 60 | 89 | 80 |
| Methionine | 17 | 16 | 18 | 31 | 21 | 7 | 11 |
| Threonine | - | <1 | - | - | <1 | - | - |
| Valine | < 1 | 1 | <1 | 1 | 1 | - | - |

### Meal Protein Quality Score per plant protein proportion

Figure 2. shows the mean (A) and distribution (B) of MPQS per plant protein proportion. MPQS decreases with increasing proportions of plant-based protein of total protein in a meal. From all n=357 fully vegan meals, no meal reached an MPQS of 100, indicating that all these meals are inadequate in some essential amino acid. The limiting EAAs of the vegan meals were (expressed as % of cases): lysine (79%), methionine (11%), cysteine (9%) and leucine (1%, **Table 5**). Inspection of these vegan meals showed that 71% of them were breakfast meals. The MPQS of all animal-rich meals frequently scored above 100, with extremes reaching 500, meaning that the consumption of the limiting essential amino acid is exceeding the requirement by 5 times.

**Figure 2.**
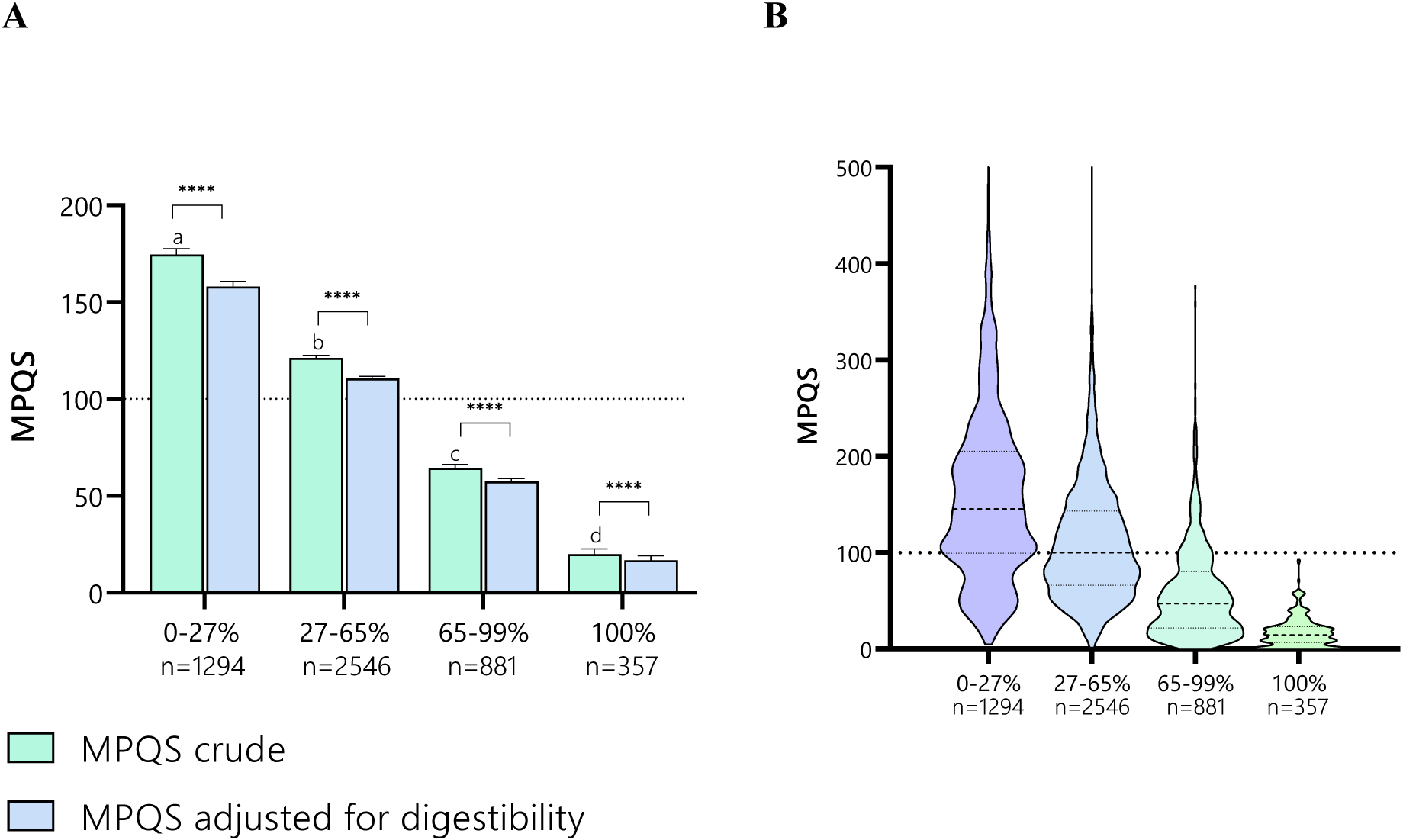
**A.** Mean Meal Protein Quality Score by plant protein proportion. This figure illustrates that meals composed entirely of plant-based proteins tend to have lower protein quality scores. Notably, within the group of meals with 100% plant protein, breakfasts constitute 71% and are generally associated with lower protein quality scores. This differentiation highlights the influence of meal type on protein quality assessments in plant-based diets. **B.** Meal Protein Quality Score Distributions by Plant Protein Proportion. This figure presents a violin plot illustrating the distribution of Meal Protein Quality Scores across different proportions of plant protein. It highlights that meals exclusively comprising plant proteins do not achieve a score of 100, whereas meals with a high proportion of animal proteins frequently exceed a score of 100, suggesting instances of protein overconsumption.

Digestibility adjustment significantly reduced MPQS in all categories of plant protein proportion. Figure 3 shows that the impact of digestibility adjustment is around 10% in the meals containing animal protein, but above 15% in the vegan meals.

**Figure 3.**
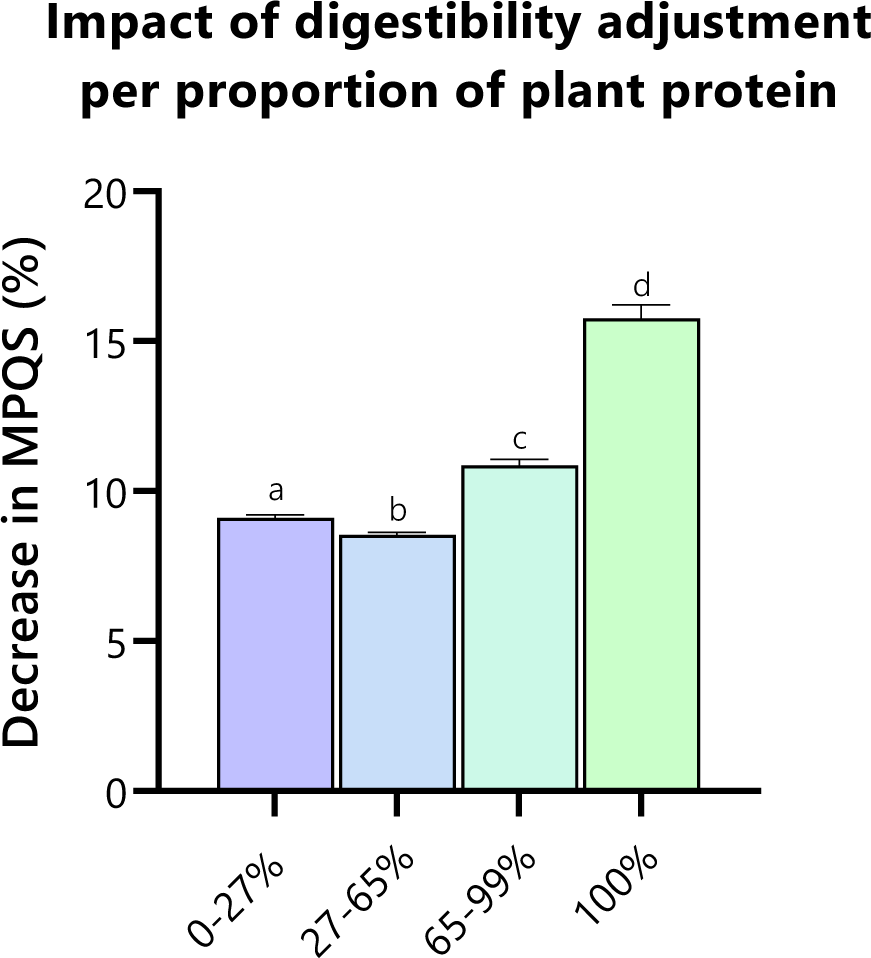
Impact of digestibility adjustment on the Meal Protein Quality Score of meals by plant protein proportion. This figure demonstrates how the inclusion of a digestibility adjustment factor affects the Meal Protein Quality Scores, particularly as the proportion of plant protein in meals increases. The relevance of digestibility adjustment is accentuated in meals with higher plant protein content, illustrating the significant role that digestibility plays in evaluating the protein quality of plant-based meals.

### Correlation of meal nutrients and MPQS

Figure 5A shows the correlation matrix between MPQS and several meal nutrient characteristics. MPQS showed to be positively correlated with amounts of total protein, plant protein, animal protein and kilocalories in a meal, while percentage of meal plant protein content showed a negative correlation. Linear regression showed that a model containing meal moment information and nutritional composition information was able to predict MPQS with an R2 of 0.77 (Figure 5B).

**Figure 5.**
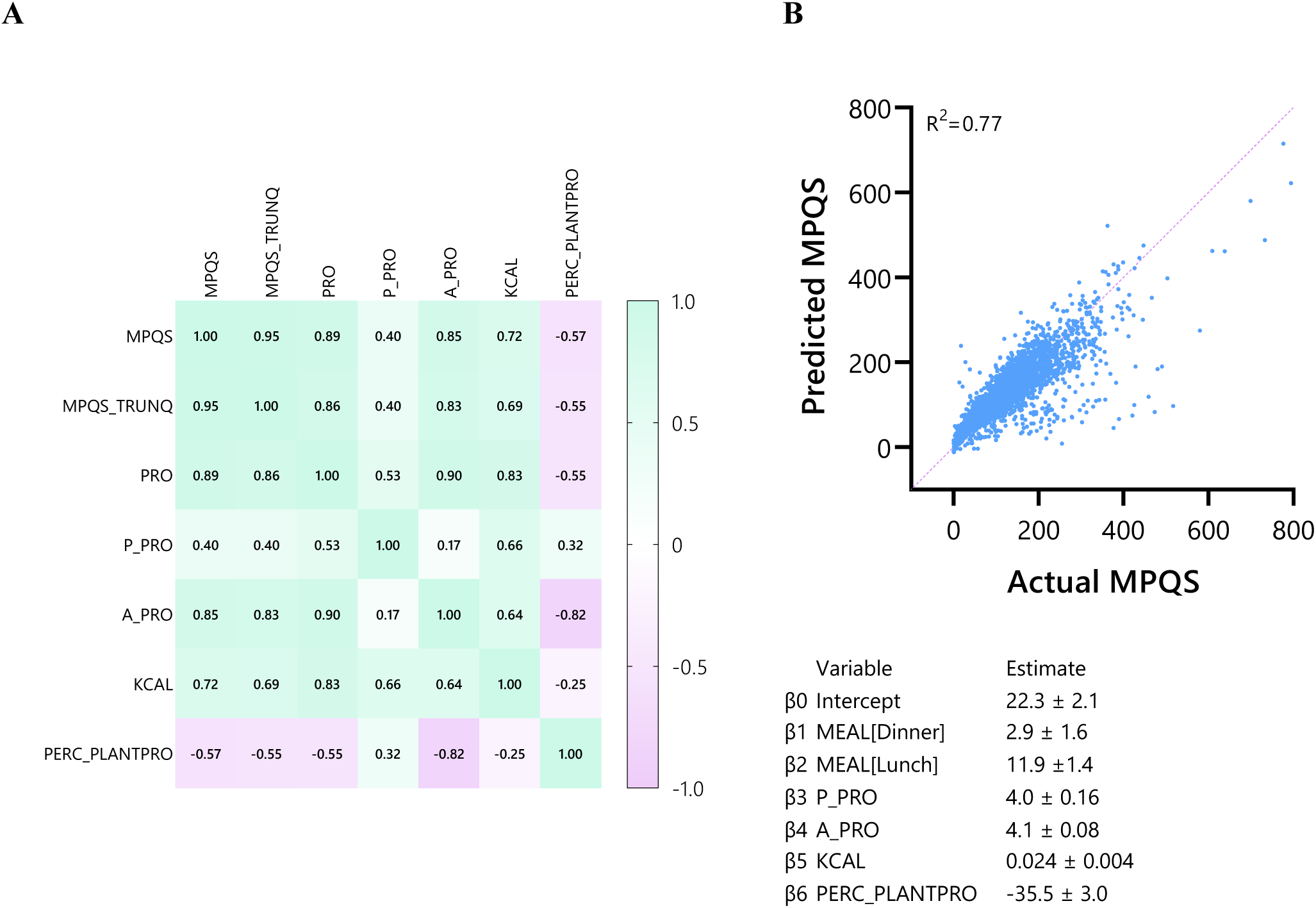
**A** Spearman’s Correlation matrix for Meal Protein Quality Score and nutritional characteristics. **B**. Predicted vs Actual Meal Protein Quality Score. This figure illustrates that the Meal Protein Quality Score can be accurately predicted using a combination of factors, including meal timing, the quantity of plant and animal proteins, total kilocalories, and the percentage of plant protein.

### Inspection of missed proteins

On average, 11.1 ± 10.9 g of protein was consumed outside of the three main meals, and thus not considered in the calculation of median daily MPQS. Out of the 1757 days that were analyzed, on 263 days (15%), the amount of protein eaten outside of the main meals exceeded 20 grams. Inspection of these 263 days revealed that the proteins mainly come from small snacks (cake, nuts, cheese etc.) and from milk. Only on 10 days (<1%), breakfast was consumed at the meal moment ‘before breakfast’ and thus missed in the analyses.

## Discussion

This paper describes the development of a new protein quality score, that combines amino acid patterns, protein digestibility, timing of combinations and protein quantity. We show how the score can be applied to make protein quality of meals insightful, and use it as a starting point of meal improvement to ensure sufficient protein quality. Additionally, we show how it can be used to quantify protein quality of individual dietary intake, in large nutritional datasets, and in real-world recipes.

Our new score, the MPQS, is of added value in the field of protein quality. Several protein quality scores already exist, such as PDCAAS an DIAAS. These scores are suitable for protein quality assessment at the product level only, whereas in practice, people eat meals, in which specific combinations of protein products are made ^3^. Other scores have been developed to address this limitation of PDCAAS and DIAAS, such as the EAA-9 score ^7^, which enables protein quality calculations similar to our developed scoring algorithm. The EAA-9 scoring framework, however, does not strictly account for the timeframe within which protein combinations must occur, nor does it establish clear amino acid requirements per specific time intervals. As a result, while the EAA-9 remains highly flexible, the numerous decisions required of the user may impede practical implementation.

Our score relies on several assumptions. First, it assumes a meal protein recommendation of 0.3 grams of perfect quality protein per kilogram body weight per meal. That number is based on several arguments. First, it seems that this protein amount per meal is sufficient to stimulate muscle protein synthesis ^32^. There are studies suggesting that an even higher dose of proteins would be ideal for older consumers ^18^, but we reason that we should err on the lower bound of the ideal protein range. That is because we optimize the protein quality within this quantity goal, meaning that we may achieve higher levels of muscle protein synthesis at lower levels of total protein quantity. Moreover, it seems inappropriate to try and find meal protein doses where muscle protein synthesis is stimulated *optimally* or *maximally* ^5^, as the latest evidence seems to point out such levels could very well exceed 100 grams of protein per meal ^29^. Finally, with three meals of 0.3 grams of optimized protein per kilogram bodyweight, and some protein intake coming from snacks, we will achieve intakes of ≥1 gram of optimized protein per kg bodyweight. There is no indisputable evidence that a daily protein intake well above 1 gram per kilogram of body weight provides health benefits for older individuals ^12^. Although moderately increased intakes do not appear to pose health risks, it is imprudent to recommend unnecessarily high intakes due to the environmental impact of protein production.

A second assumption is that combinations of protein sources should be made within one meal. The current state of research does not clearly point to a specific time frame in which protein sources should be combined to complement each other’s amino acid profiles ^9^. Some argue that this should be ideally done within a meal, especially on lower protein diets or when specific essential amino acids are limiting on sequential days ^15^, while others assume protein combinations can be made over a whole day ^17^. Although there are no strict storage pools for free amino acids, recent work from Pinckaers et al. ^20, 21^ would suggest that the body is capable of correcting for a unbalanced amino acid composition at large doses of protein. While their results suggest that lacking amino acids may have been corrected for by plasma pools, the precise mechanism is insufficiently elucidated. In animals, so-called labile storage proteins have been discussed as another potential mechanism, supporting this amino acid buffering feature ^24, 27^. However, the capacity to correct for an amino acid disbalance was limited, as the released EAAs were still greater when comparing whey to gelatine ^27^. Consequently, excessive amino acid disbalance will likely still result in suboptimal utilization of the ingested proteins, while the labile storage proteins have so far also not been identified in humans. Interestingly, a recent study explored the impact of daily supplementation with 50 grams of whey protein, pea protein, or collagen protein on muscle protein synthesis in older adults over a week ^16^. The findings revealed that unlike whey and pea proteins, collagen protein did not enhance muscle protein synthesis. The authors suspect that the low leucine content in collagen protein may be responsible for its inability to promote muscle protein synthesis. However, it is also plausible that the complete absence of the essential amino acid tryptophan in collagen (resulting in an MPQS of 0) contributes to its failure to stimulate muscle protein synthesis, thereby supporting the notion that a meal must contain all essential amino acids in the proper balance to effectively stimulate protein synthesis. Until the debate around the time window of protein complementation is settled, we err on the strict side and assume the meal moment is the time window in which protein sources should be combined. If needed, our scoring mechanism can be adapted into a Daily Protein Quality Score, by inputting all daily intake and using daily amino acid RDAs, or a combination of a daily protein target (such as 0.8 or 1.2 g/kg/d) with an amino acid reference pattern.

Thirdly, we assume that the amino acid reference patterns recommended by FAO and WHO represents ideal quantities for older adults. These reference patterns are established for the total population, while older adults may benefit from higher amounts of certain amino acids, such as leucine ^26^. Moreover, where other scoring algorithms combine methionine and cysteine by simply using their sum, we use a more complex assumption where only methionine consumed above the recommendation will be converted into cysteine, and never the other way around ^25^. That is justified based on metabolic possibilities. However, some studies suggest that the methyl donating capacities of the sulfuric amino acids are the main driver of their physiological role, suggesting that using their sum could be appropriate after all ^8^.

In this study, we observed that more plant-based meals are often of lower quality. That is in line with many previous observations. Importantly, this study did not specifically include vegetarians and vegans, who may be experienced in making better combinations and thus achieve higher protein quality in their predominantly plant-based meals. Moreso, our study sample tended to consume the majority of their animal proteins at dinner, and the lowest at breakfast. In our observations, over 70% of the vegan meals were breakfasts. Breakfasts are known to be the lowest scoring meals in term of protein quantity ^28^ and quality, therefore confounding the relationship between the plant protein proportion of a meal and its protein quality.

Moreover, dividing the study population in median MPQS above or below 100 revealed that the high MPQS group had a 15% higher energy intake and a 25% higher protein intake, suggesting generally higher consumption levels, particularly of protein. Interestingly, the BMI in the high MPQS group was 7% lower compared to the low MPQS group. This disparity suggests potential underreporting of energy intake, yet it does not account for the notably higher protein consumption observed in the high MPQS group. Possibly, a higher protein quality intake has an effect on body composition by a larger stimulation of fat free mass synthesis, but that hypothesis needs to be investigated.

In conclusion, we present a new scoring algorithm that enables the calculation of protein quality of meals and recipes: the Meal Protein Quality Score. This score is much needed to guide the protein transition towards plant-rich meals of high quality, so that vulnerable populations such as older adults and patients can safely transit to more plant-based diets as well. We show that the MPQS can be used to qualify and improve recipes, to calculate individual protein quality intake per meal, and to calculate protein quality of all meals in large epidemiological studies.

## Data Availability

All data produced in the present study are available upon reasonable request to the authors

## Abbreviations

EAA: Essential Amino Acid
DIAAS: Digestible Indispensable Amino Acid Score
MPQS: Meal Protein Quality Score
PDCAAS: Protein Digestibility-Corrected Amino Acid Score

## Acknowledgements

PG, SHC, MT, WR, LdG, IT designed research; PG conducted research, analyzed data and wrote the paper; SHC, MT, WR, IT, LdG provided essential materials.

## The authors declare no conflict of interest

Data described in the manuscript, code book, and analytic code will be made available upon request pending application and approval.

